# Association of magnesium depletion score with cardiovascular disease and its association with longitudinal mortality in cardiovascular disease patients

**DOI:** 10.1101/2023.03.06.23286882

**Authors:** Liu Ye, Cheng Zhang, Qin Duan, Yue Shao, Jianzhong Zhou

## Abstract

**Background:** Dietary magnesium and serum magnesium play an important part in cardiovascular disease (CVD). However, the correlation between magnesium depletion score (MDS) and the development and CVD prognosis remains unclear. This analysis examines the cross-sectional relationship between MDS and CVD, and the longitudinal correlation between MDS and all-cause and cardiovascular mortality in CVD individuals.

**Methods:** In all, 42,711 individuals were selected from the National Health and Nutrition Examination Survey, including 5,015 subjects with CVD. The correlation between MDS and total and individual CVD was examined using the survey-weighted multiple logistic regression analysis. Among 5,011 CVD patients, 2,285 and 927 participants were recorded with all-cause and cardiovascular deaths, respectively. We applied survey-weighted Cox proportional hazards regression analyses to investigate the impact of MDS on the mortality of CVD individuals.

**Results:** CVD group had higher MDS levels than the non-CVD groups. After controlling all confounding factors, individuals with MDS scored 1-2, and ≥3 had higher odds of total CVD and specific CVD than those with MDS scored 0. The relationship between MDS and total CVD was stable and significant in all subgroups. The fully adjusted Cox regression model presented that the risk of all-cause and cardiovascular deaths increased 2.41 and 2.00 times in participants with MDS≥3 compared to those with MDS scored 0.

**Conclusions:** MDS is a vital risk factor for CVD’s prevalence and all-cause and cardiovascular deaths.

**What is new?:**

- MDS is a significant positive correlation with CVD’s prevalence in US adults.
- High MDS markedly elevated the risk of all-cause and cardiovascular deaths among participants with CVD.

**What are the clinical implications?:**

- MDS can be used to evaluate the prevention and prognosis of cardiovascular diseases.

## 1. Introduction

Cardiovascular disease (CVD) is a broad term, including heart failure, coronary heart disease (CHD), angina, heart attack, and stroke. It is recognized as the leading cause of worldwide deaths, approximately 30% annually^1^. Besides traditional cardiovascular risk factors, mineral disorders can increase CVD occurrence^2^.

Magnesium, an essential and abundant mineral, is involved in numerous metabolic reactions in the body, including ATP production, maintaining normal mitochondrial function, glycolysis, and protein and DNA synthesis^3, 4^. Magnesium has also been crucial in regulating blood pressure, myocardial metabolism, cardiac contractility and excitability, and vascular tone^4, 5^. Hypomagnesemia has been associated with elevated risk and adverse prognosis in individuals with diabetes, heart failure, coronary artery disease, and stroke^6-10^. Increased dietary magnesium intake reduced the risk of diabetes, stroke, metabolic syndrome, and hypertension^11-14^.

Previous research primarily concerned about the impacts of dietary magnesium and serum magnesium on CVD, ignoring magnesium deficiency. The magnesium depletion score (MDS) is a comprehensive scoring tool for evaluating magnesium deficiency status. A higher MDS suggests a greater severity of magnesium deficiency. Evidence from Fan et al. study showed that individuals with high MDS had elevated all-cause and cardiovascular death risk^15^. Jian et al. provided evidence that elevated MDS was related to a higher abdominal aortic calcification (AAC) prevalence^16^. However, research on MDS and CVD incidence is limited. Additionally, given that CVD substantially shortens life expectancy, investigating the correlation between MDS with all-cause and cardiovascular deaths among CVD patients would have implications for tertiary prevention.

We conducted a cross-sectional and longitudinal study based on the National Health and Nutrition Examination Survey (NHANES) to solve this problem. This paper dedicated to explore whether 1) MDS is cross-sectionally related to a higher CVD prevalence; and 2) MDS is longitudinally correlated with elevated risks of all-cause and cardiovascular deaths among participants with CVD.

## 2. Materials and methods

### 2.1. Study population

NHANES is a nationwide cross-sectional survey that reflects the health and nutrition status of the general population in the United States using a multistage probability sampling design^17^. NHANES has acquired written informed consent from each participant. Approval was obtained from the Research Ethics Review Board of the National Center for Health Statistics (NCHS). Therefore, no additional ethical approval was needed for this analysis.

Part I: We assessed 101,316 participants from ten continuous cycles (1999-2018) of NHANES. After excluding individuals younger than 20 years, pregnant, and lacking available MDS and CVD information, 42,711 respondents were selected. Among those, 5,015 individuals self-reported physician diagnosis of CVD. The cross-sectional correlation between MDS and CVD was explored (Figure 1).

**Figure 1.**
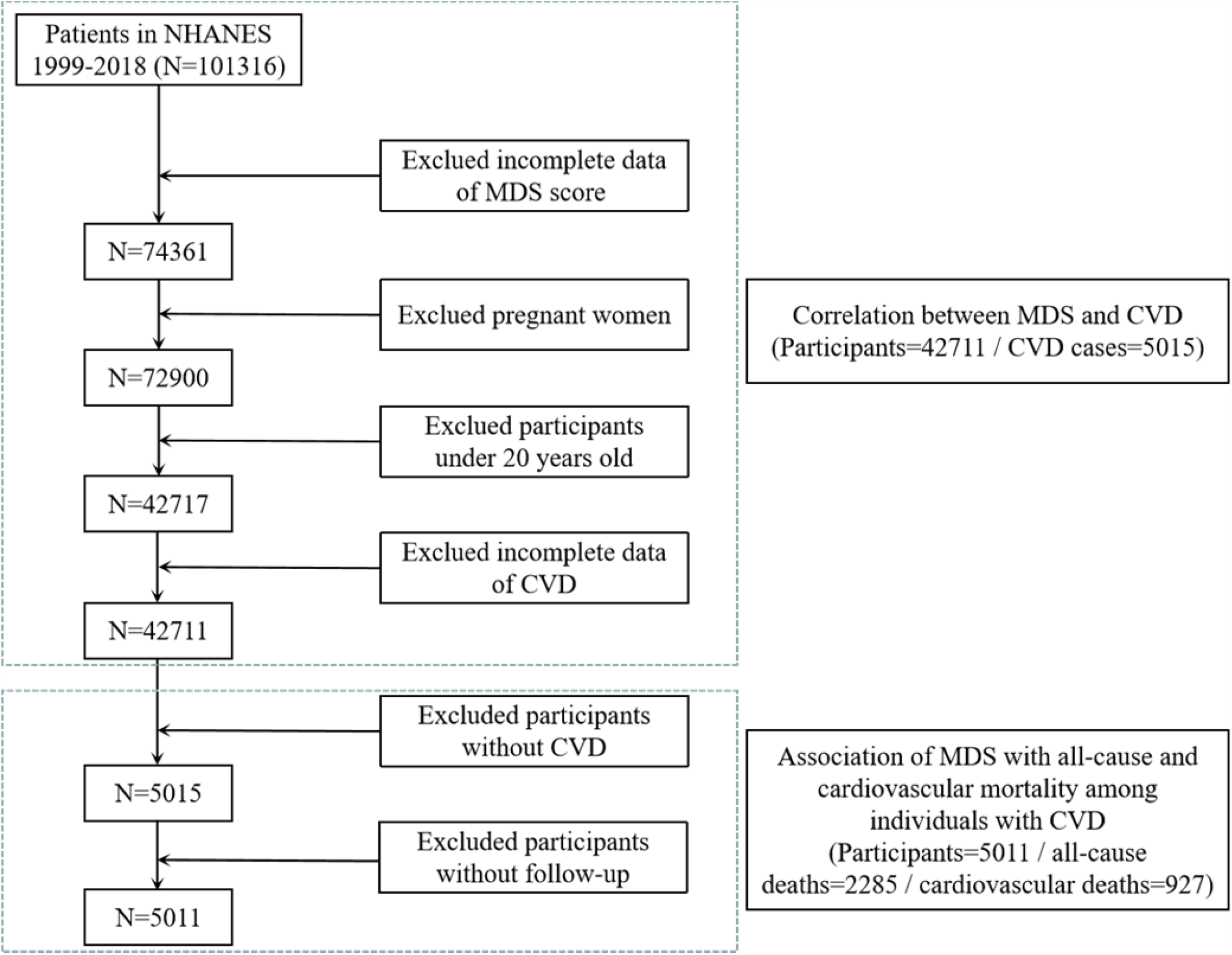
Flowchart of the sample selection from NHANES 1999–2018.

Part II: Next, we excluded 4 individuals who lost to follow-up. Among 5011 individuals with CVD, 2285 and 927 participants were recorded with all-cause and cardiovascular deaths, respectively, until December 31, 2019. The longitudinal relationship between MDS and mortality was investigated among CVD patients (Figure 1).

### 2.2. Assessment of outcomes

Part I: CVD was ascertained using five self-reported CVD subtypes, containing congestive heart failure (CHF), CHD, angina, heart attack, or stroke. Each respondent was asked, “ Has a doctor or other health professional ever told you that you have CHF/CHD/angina pectoris/heart attack/stroke?” Answering “ yes” to any above questions was regarded as having CVD.

Part II: The primary outcomes were death due to any cause and CVD (codes I00-I09,I11,I13, I20– I51,and I60–I69). The cause and time of death were ascertained by NCHS Linked Mortality Files updated through December 31, 2019.

### 2.3. MDS assessment

The MDS was calculated as the sum of the following four scores: 1) current use of diuretics was scored 1 point; 2) current use of PPI was scored 1 point; 3) 60 mL/(min 1.73 m^2^) ≤ eGFR < 90 mL/(min 1.73 m^2^) was scored 1 point, or eGFR <60ml/(min 1.73 m^2^) was scored 2 points. 4) Heavy drinkers (>1 drink/d for females and >2 drinks/d for males) was scored 1 point.

### 2.4. Covariates

The information contained sociodemographic characteristics [age, gender, marital status, ethnicity, educational attainment, family poverty-to-income ratio, and body mass index (BMI)], lifestyle behaviors (physical activity, smoking status, and alcohol consumption), dietary factors (intakes of total energy and magnesium), comorbidities (hypertension, diabetes, and hyperlipidemia), and laboratory data (high-density cholesterol and total cholesterol). The dietary data was acquired via 24 h dietary recall surveys. Diabetes was considered by self-reported diagnosis, using diabetes medications, hemoglobin A1c ≥ 6.5%, or fasting plasma glucose ≥ 126 mg/dl. Hypertension was diagnosed by self-reported diagnosis, using anti-hypertension medications, or systolic/diastolic blood pressure ≥ 140/90 mmHg. Hyperlipidemia was determined by self-reported diagnosis, using anti-hyperlipidemia drugs, triglyceride ≥ 200 mg/dL, high-density lipoprotein cholesterol < 40 mg/dL, or low-density lipoprotein cholesterol ≥ 130 mg/dL.

### 2.5. Statistical analysis

The data was processed in accordance with NHANES analytical guidelines. All analyses used appropriate sample weights and strata due to the complex sampling design of NHANES^18^. Continuous variables were summarized as the weighted mean ± standard error and examined using the weighted linear regression. Categorical data were given in the form of the weighted percentages, and comparison between groups was investigated using the weighted Chi-square test. The relationship between MDS and total CVD was examined using survey-weighted multiple logistic regression analyses in three different models, with age, sex, and ethnicity adjusted in Model I; remaining sociodemographic characteristics, lifestyle behaviors, and comorbidities added in Model II; and dietary factors and laboratory data added in Model III. Survey-weighted multiple logistic regression analyses were carried out to clarify the correlation between MDS and the five individual CVD prevalences (CHF, CHD, angina, heart attack and stroke).

Stratification analysis was implemented to investigate whether the correlation between MDS and CVD was sustained across the different subgroups classified using age, gender, smoking status, BMI, magnesium intake, and complications.

We performed weighted Kaplan-Meier curves with the log-rank tests for cumulative survival differences across different MDS scores. Survey-weighted Cox proportional hazards regression analyses were employed to examine the relationship between MDS and all causes and cardiovascular mortality in the multivariate model, where three models were created consistent with the above (survey-weighted multiple logistic regression analyses). We employed all statistical analyses using R software (version 3.6.3). *P* values < 0.05 were considered significant.

## 3. Results

### 3.1. The relationship between MDS and CVD

This study selected 42,711 individuals from NHANES, including 5,015 participants with CVD. Table 1 compares the main characteristics of subjects based on with or without CVD. CVD group had higher MDS levels compared to non-CVD groups. Subjects with CVD were older, more likely to be male, Non-Hispanic white, had higher BMI, former smokers and drinkers, higher hypertension, diabetes, and hyperlipidemia incidence, and lower high-density cholesterol levels. They were more likely to never exercise, have lower education levels and family income, and consume less energy and magnesium. However, total cholesterol levels was significantly lower in participants with CVD, which might be due to the use of lipid-lowering drugs.

**Table 1.**
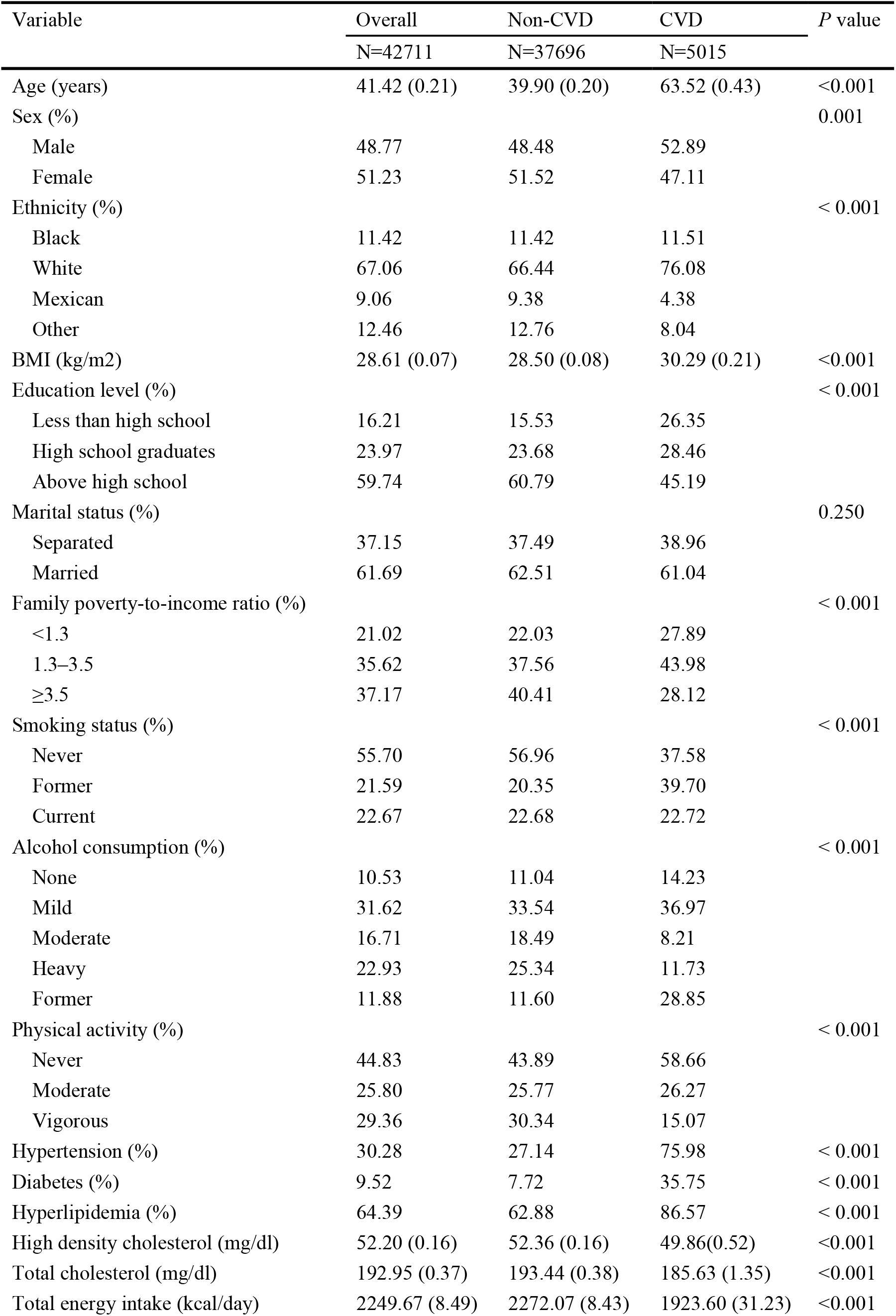
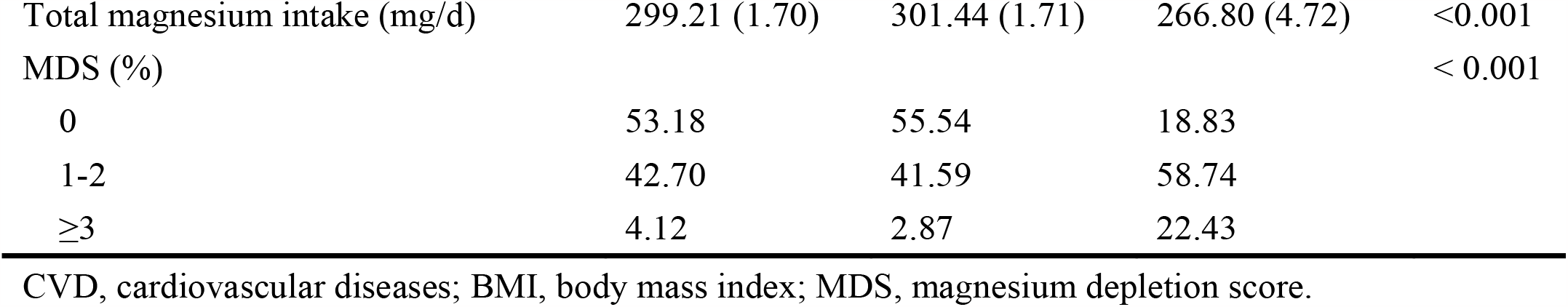
Participant characteristics in NHANES 1999–2018, weighted.

Table 2 presents the correlation between MDS and the total CVD risk using the survey-weighted multivariate logistic regression model. After controlling all confounding factors (Model III), individuals with MDS scored 1-2, and ≥3 had 2.02 (1.67, 2.45), and 3.76 (2.89, 4.87) folds the risk of CVD, respectively, (*P* for trend < 0.001) compared to those with MDS=0.

**Table 2.** The relationship between MDS and the prevalence of total CVD (Participants=42,711 / CVD cases=5,015).

|  | Model I |  | Model II |  | Model III |  |
| --- | --- | --- | --- | --- | --- | --- |
|  | OR (95% CI) | *P* | OR (95% CI) | *P* | OR (95% CI) | *P* |
| MDS |  |  |  |  |  |  |
| 0 | Reference |  | Reference |  | Reference |  |
| 1-2 | 2.34 (1.96, 2.80) | <0.001 | 1.89 (1.57, 2.26) | <0.001 | 2.02 (1.67, 2.45) | <0.001 |
| ≥3 | 6.29 (5.02, 7.88) | <0.001 | 3.61 (2.82, 4.63) | <0.001 | 3.76 (2.89, 4.87) | <0.001 |
| *P* for trend | <0.001 |  | <0.001 |  | <0.001 |  |

MDS, magnesium depletion score; CVD, cardiovascular diseases; OR, odds ratio; 95% CI, 95% confidence interval.
Model I adjusted for age, sex, and ethnicity.
Model II adjusted for model I + education level, marital status, family poverty-to-income ratio, smoking status, alcohol consumption, physical activity, BMI, hypertension, diabetes, and hyperlipidemia.
Model III adjusted for model II +high density cholesterol, total cholesterol, energy intake and magnesium intake.

The correlation between MDS and the risk of five specific CVD revealed a similar trends in the Table 3. After adjusting for all covariates, participants with MDS ≥ 3 were correlated with a greater prevalence of CHF [OR (95% CI): 7.45 (4.62, 12.00), *P* for trend <0.001], CHD [OR (95% CI): 5.24 (3.36, 8.19), *P* for trend <0.001], angina [OR (95% CI): 2.22 (1.41, 3.50), *P* for trend =0.014], heart attack [OR (95% CI): 3.76 (2.43, 5.82), *P* for trend <0.001], and stroke [OR (95% CI): 3.09 (2.13, 4.47), *P* for trend <0.001] relative to those with MDS scored 0.

**Table 3.** Adjusted odds ratios for correlations between MDS and individual CVDs (Participants=42,711 / CVD cases=5,015).

|  | Congestive heart failure | Coronary heart disease | Angina pectoris | Heart attack | Stroke |
| --- | --- | --- | --- | --- | --- |
|  | OR (95% CI) | OR (95% CI) | OR (95% CI) | OR (95% CI) | OR (95% CI) |
| MDS |  |  |  |  |  |
| 0 | Reference | Reference | Reference | Reference | Reference |
| 1-2 | 2.36 (1.46, 3.82)*** | 2.76 (1.90, 3.99)*** | 1.52 (1.07, 2.18)* | 2.38 (1.65, 3.42)*** | 1.91 (1.41, 2.58)*** |
| ≥3 | 7.45 (4.62, 12.00)*** | 5.24 (3.36, 8.19)*** | 2.22 (1.41, 3.50)*** | 3.76 (2.43, 5.82)*** | 3.09 (2.13, 4.47)*** |
| *P* for trend | <0.001 | <0.001 | 0.014 | <0.001 | <0.001 |

\* $P<0.05$ ; \*\* $P<0.01$ ; \*\*\* $P<0.001$ .
MDS, magnesium depletion score; CVD, cardiovascular diseases; OR, odds ratio; 95% CI, 95% confidence interval.
Survey-weighted multiple logistic adjusted for age, sex, ethnicity, education level, marital status, family poverty-to-income ratio, smoking status, alcohol consumption, physical activity, BMI, hypertension, diabetes, hyperlipidemia, high density cholesterol, total cholesterol, energy intake and magnesium intake.

Stratified analyses indicated stable and consistent associations between MDS and increased risk of CVD in all subgroups (Table 4). Additionally, individuals without hyperlipidemia had a stronger relationship between MDS and CVD than those with hyperlipidemia (P_interaction_ =0.042).

**Table 4.** Subgroup analysis for the association between MDS and CVD (Participants=42,711 / CVD cases=5,015).

|  | MDS |  |  | *P* for trend | *P* for interaction |
| --- | --- | --- | --- | --- | --- |
|  | 0<br>OR (95% CI) | 1-2<br>OR (95% CI) | ≥3<br>OR (95% CI) |  |  |
| Magnesium intake |  |  |  |  | 0.302 |
| 0-262 mg | Reference | 2.35 (1.80, 3.07) | 4.42 (3.08, 6.35) | <0.001 |  |
| >262 mg | Reference | 1.67 (1.24, 2.25) | 3.05 (2.01, 4.63) | <0.001 |  |
| Age |  |  |  |  | 0.604 |
| <65 years | Reference | 1.84 (1.50, 2.25) | 3.52 (2.35, 5.27) | <0.001 |  |
| ≥65 years | Reference | 1.87 (1.08, 3.24) | 3.73 (2.10, 6.63) | <0.001 |  |
| Sex |  |  |  |  | 0.188 |
| Male | Reference | 2.13 (1.60, 2.83) | 4.20 (2.80, 6.29) | <0.001 |  |
| Female | Reference | 1.90 (1.42, 2.55) | 3.53 (2.45, 5.11) | <0.001 |  |
| Smoking status |  |  |  |  | 0.735 |
| Never | Reference | 1.84 (1.29, 2.62) | 3.11 (1.91, 5.07) | <0.001 |  |
| Former | Reference | 1.99 (1.36, 2.91) | 3.98 (2.44, 6.51) | <0.001 |  |
| Current | Reference | 2.16 (1.61, 2.90) | 4.66 (2.44, 8.92) | 0.003 |  |
| BMI |  |  |  |  | 0.219 |
| <25 kg/m<sup>2</sup> | Reference | 1.88 (1.23, 2.87) | 2.90 (1.62, 5.19) | 0.019 |  |
| 25–30 kg/m<sup>2</sup> | Reference | 2.23 (1.45, 3.41) | 5.35 (3.26, 8.80) | <0.001 |  |
| ≥30 kg/m<sup>2</sup> | Reference | 1.94 (1.47, 2.57) | 3.24 (2.25, 4.66) | <0.001 |  |
| Hypertension |  |  |  |  | 0.081 |
| Yes | Reference | 1.62 (1.18, 2.22) | 3.91 (1.82, 8.40) | 0.010 |  |
| No | Reference | 2.11 (1.61, 2.76) | 4.01 (2.89, 5.56) | <0.001 |  |
| Diabetes |  |  |  |  | 0.106 |
| Yes | Reference | 1.94 (1.51, 2.49) | 3.71 (2.71, 5.08) | <0.001 |  |
| No | Reference | 2.25 (1.55, 3.27) | 4.17 (2.62, 6.64) | <0.001 |  |
| Hyperlipidemia |  |  |  |  | 0.042 |
| Yes | Reference | 1.30 (0.78, 2.16) | 3.62 (1.87, 6.98) | <0.001 |  |
| No | Reference | 2.21 (1.80, 2.73) | 3.99 (3.03, 5.26) | <0.001 |  |

MDS, magnesium depletion score; CVD, cardiovascular diseases; BMI, body mass index; OR, odds ratio; 95% CI, 95% confidence interval.

### 3.2. The relationship of MDS with mortality among individuals with CVD

A total of 5,011 CVD subjects had available follow-up data. This study identified 2,285 all-cause deaths and 927 cardiovascular deaths over 81 months of the median follow-up period. The Kaplan-Meier curves depicted the survival difference between patients with different MDS scores. For all-cause deaths, participants with MDS ≥ 3 had the lowest survival probability, whereas those with MDS = 0 had the highest survival probability (log-rank *P* < 0.001, Figure 2A). Figure 2B demonstrates a similar pattern in cardiovascular deaths (log-rank *P* < 0.001).

**Figure 2.**
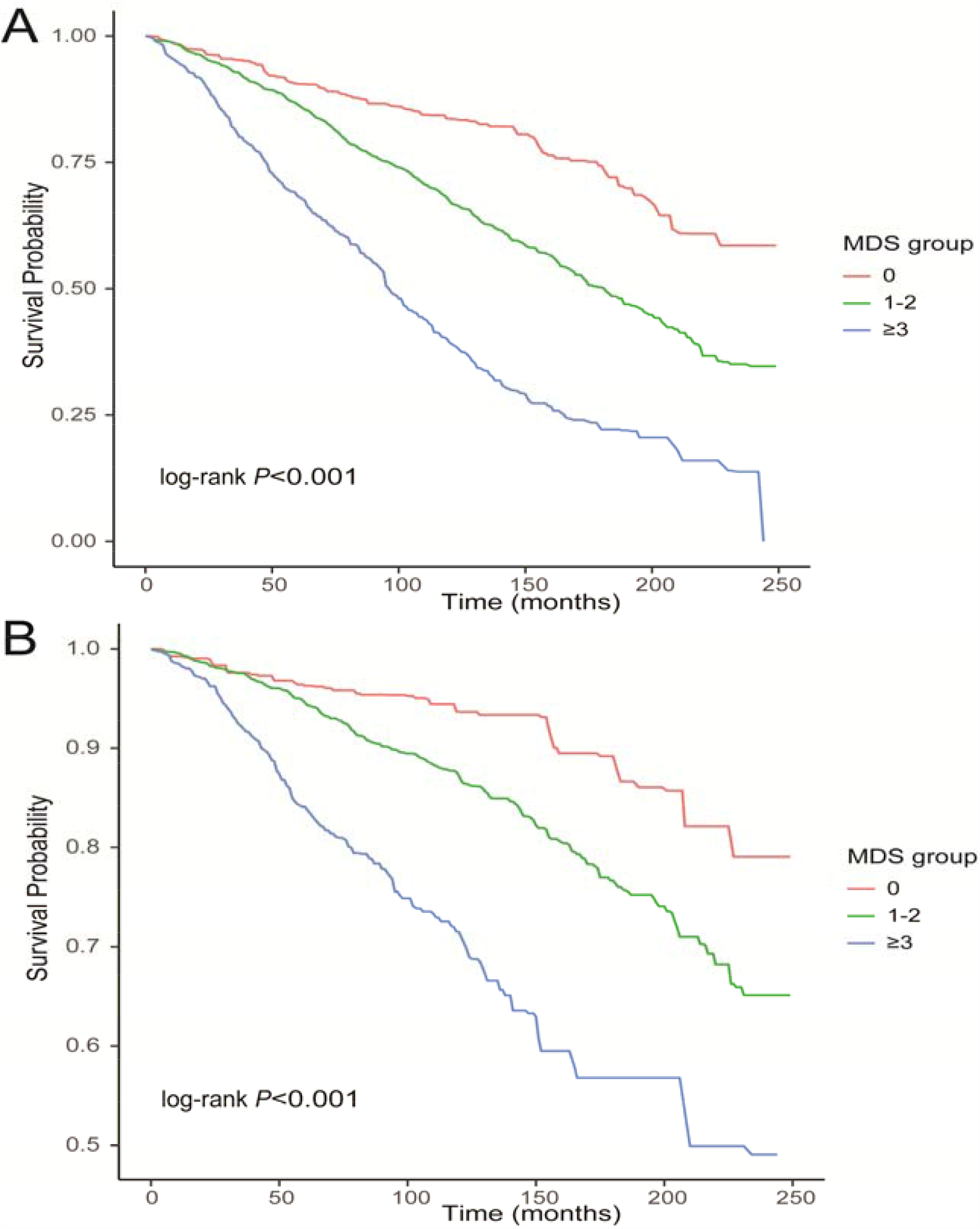
Kaplan-Meier curves were used to present the relationship of the MDS with all-cause (A) and cardiovascular mortality (B).

Table 5 reveals the relationship of MDS with all-cause and cardiovascular deaths using three different Cox regression models. After fully controlling for confounder variables (Model III), the risk of all-cause deaths increased 2.41 times in participants with MDS ≥ 3 [HR (95% CI): 2.41 (1.70, 3.40), P for trend < 0.001] compared to those with MDS scored 0. Similarly, subjects with MDS ≥ 3 had a higher risk of cardiovascular deaths than those with MDS = 0 [HR (95% CI): 2.00 (1.10, 3.62), P for trend < 0.001].

**Table 5.** Correlation of MDS with the risk of all-cause and cardiovascular mortality among 5,011 participants with CVD.

|  | MDS |  |  | *P* for trend |
| --- | --- | --- | --- | --- |
|  | 0 | 1-2 | ≥3 |  |
|  | HR (95% CI) | HR (95% CI) | HR (95% CI) |  |
| All-cause mortality |  |  |  |  |
| Number of deaths | 204/763 | 1,325/3,043 | 756/1,205 |  |
| Model I | Reference | 1.22 (0.94, 1.58) | 2.27 (1.75, 2.94)*** | <0.001 |
| Model II | Reference | 1.25 (0.94, 1.67) | 2.20 (1.62, 2.99)*** | <0.001 |
| Model III | Reference | 1.35 (0.97, 1.88) | 2.41 (1.70, 3.40)*** | <0.001 |
| CVD mortality |  |  |  |  |
| Number of deaths | 61/763 | 536/3,043 | 330/1,205 |  |
| Model I | Reference | 1.05 (0.70, 1.60) | 2.00 (1.30, 3.08)** | <0.001 |
| Model II | Reference | 1.12 (0.67, 1.89) | 2.03 (1.18, 3.50)* | <0.001 |
| Model III | Reference | 1.11 (0.64, 1.96) | 2.00 (1.10, 3.62)* | <0.001 |

\* $P<0.05$ ; \*\* $P<0.01$ ; \*\*\* $P<0.001$ .
MDS, magnesium depletion score; CVD, cardiovascular diseases; HR, hazard ratios; 95% CI, 95% confidence interval.
Model I adjusted for age, sex, and ethnicity.
Model II adjusted for model I + education level, marital status, family poverty-to-income ratio, smoking status, alcohol consumption, physical activity, BMI, hypertension, diabetes, and hyperlipidemia.
Model III adjusted for model II + high density cholesterol, total cholesterol, energy intake and magnesium intake.

## 4. Discussion

This finding revealed a significant positive correlation between MDS and CVD in US adults. The relationship between MDS and CVD remained significant in all subgroups, suggesting that this correlation may be applicable to different populations. Additionally, high MDS markedly elevated the risk of all-cause and cardiovascular deaths among participants with CVD.

Serum magnesium is the most commonly used approach to evaluate magnesium status in clinical practice. Several studies have reported hypomagnesaemia as a significant risk factor for CVD ^7-9, 19, 20^. However, serum magnesium cannot reflect the actual status of whole body magnesium levels. The serum magnesium level is typically within the normal reference range, especially in the case of chronic magnesium deficiency^21, 22^. The serum contains 0.3% of the body’s total magnesium; and the rest is primarily stored in bones, muscles, and soft tissue^23^. Urinary magnesium level is another way to assess magnesium status. A 24-h urinary magnesium/creatinine ratio has been proved to be negatively correlated with cardiovascular risk factors^24^. Nevertheless, urinary magnesium is easily affected by dietary intake, proton pump inhibitor (PPI), diuretic, and renal function^21, 25^. Besides, the evaluation experiment is complex specially for older people, because it requires 24 h urine collection^26^. The magnesium tolerance test (MTT) is regarded as a gold standard for evaluating the magnesium status of the body, but it is unpractical and difficult to apply broadly since it requires first 24 h urine collection, intravenous magnesium infusion, and then second 24 h urine collection^27^. Moreover, MTT is unsuitable for patients with impaired renal function.

MDS has been demonstrated as a good indicator for predicting magnesium deficiency validated by MTT. It combines four risk factors affecting magnesium reabsorption in the United States population, including alcohol consumption, diuretic, PPI, and kidney function^15^. Alcohol is considered as a magnesium diuretic, which rapidly increases magnesium excretion by causing proximal renal tubular dysfunction, and it even occurs in patients with a negative magnesium balance^28^. Similarly, diuretics can promote magnesium excretion^29^. PPI affects intestinal absorption of magnesium by down-regulating TRPM6 activity^30^. Magnesium homeostasis is mainly maintained through the kidney, where more than 80% of serum magnesium is reabsorbed^31^. Therefore, MDS is a simple and reliable tool to evaluate magnesium deficiency. In this context, we investigated the relationship between MDS and the occurrence and prognosis of CVD.

This study focuses on magnesium deficiency, and reveals a positive correlation between MDS and total and individual CVD risk. Moreover, this correlation was not affected by magnesium intake. This finding contradicted a prior study, that revealed a significant relationship between MDS and AAC only in patients with lower magnesium intake^16^.

The previous study primarily concentrated on the predictive significance of MDS on all-cause and cardiovascular mortality in the general population^15^. However, the fact is that CVD patients have a shorter life expectancy. Considering that MDS correlated with a greater risk of all-cause and cardiovascular deaths among participants with CVD in this analysis, timely intervention in CVD patients with high MDS might help to reduce death risk.

Magnesium deficiency is common in older people. Possible reasons include decreased magnesium intake and absorption, and increased excretion due to drug use^32, 33^. Our subgroup analysis revealed that the correlation between MDS and CVD persisted among different age groups individuals, which suggested that the relationship may not influenced by the age.

Although a large sample size and long-term NHANES follow-up supported our research, we must acknowledge some limitations. First, the cross-sectional study design cannot definitively determine the causation between MDS and CVD. Second, we did not compare MSD to serum magnesium due to the lack of serum magnesium in NHANES. Third, as a study based on the United stated population, ethnic differences prevent our results from being extended to the general population. Finally, although a multivariate analysis was performed, we cannot rule out the possibility of a selection bias affecting our results.

## 5. Conclusion

This current study indicated a positive correlation between MDS and CVD prevalence. Besides, high MDS elevated all-cause and cardiovascular mortality risk among individuals with CVD.

## Data Availability

All data can be obtained from NHANES official (website https://www.cdc.gov/nchs/nhanes/index.htm).

